# Identifying Women with Post-Delivery Posttraumatic Stress Disorder using Natural Language Processing of Personal Childbirth Narratives

**DOI:** 10.1101/2022.08.30.22279394

**Authors:** Alon Bartal, Kathleen M. Jagodnik, Sabrina J. Chan, Mrithula S. Babu, Sharon Dekel

## Abstract

**Background:** Maternal mental disorders are considered a leading complication of childbirth and a common contributor to maternal death. In addition to undermining maternal welfare, untreated postpartum psychopathology can result in child emotional and physical neglect, and associated significant pediatric health costs. Some women may experience a traumatic childbirth and develop posttraumatic stress disorder (PTSD) symptoms following delivery (CB-PTSD). Although women are routinely screened for postpartum depression in the U.S., there is no recommended protocol to inform the identification of women who are likely to experience CB-PTSD. Advancements in computational methods of free text has shown promise in informing diagnosis of psychiatric conditions. Although the language in narratives of stressful events has been associated with post-trauma outcomes, whether the narratives of childbirth processed via machine learning can be useful for CB-PTSD screening is unknown.

**Objective:** This study examined the utility of written narrative accounts of personal childbirth experience for the identification of women with provisional CB-PTSD. To this end, we developed a model based on natural language processing (NLP) and machine learning (ML) algorithms to identify CB-PTSD via classification of birth narratives.

**Study Design:** A total of 1,127 eligible postpartum women who enrolled in a study survey during the COVID-19 era provided short written childbirth narrative accounts in which they were instructed to focus on the most distressing aspects of their childbirth experience. They also completed a PTSD symptom screen to determine provisional CB-PTSD. After exclusion criteria were applied, data from 995 participants was analyzed. An ML-based Sentence-Transformer NLP model was used to represent narratives as vectors that served as inputs for a neural network ML model developed in this study to identify participants with provisional CB-PTSD.

**Results:** The ML model derived from NLP of childbirth narratives achieved good performance: AUC 0.75, F1-score 0.76, sensitivity 0.8, and specificity 0.70. Moreover, women with provisional CB-PTSD generated longer narratives (t-test results: *t=2*.*30, p=0*.*02*) and used more negative emotional expressions (Wilcoxon test: ‘sadness’: *p=8*.*90e-*^*04*^, *W=31,017*; ‘anger’: *p=1*.*32e-*^*02*^, *W=35,005*.*50*) and death-related words (Wilcoxon test: *p=3*.*48e-*^*05*^, *W=34,538*) in describing their childbirth experience than those with no CB-PTSD.

**Conclusions:** This study provides proof of concept that personal childbirth narrative accounts generated in the early postpartum period and analyzed via advanced computational methods can detect with relatively high accuracy women who are likely to endorse CB-PTSD and those at low risk. This suggests that birth narratives could be promising for informing low-cost, non-invasive tools for maternal mental health screening, and more research that utilizes ML to predict early signs of maternal psychiatric morbidity is warranted.

## Introduction

Approximately 140 million women give birth every year worldwide, and among them, an estimated one-third experience a highly stressful, potentially traumatic birth.^1-4^ The emotional toll of this exposure to trauma can result in a mental illness formally recognized as posttraumatic stress disorder (PTSD) that has been traditionally associated with war, combat, and serious sexual assault.^5^ However, the possibility that childbirth-related trauma could be significant enough to cause PTSD symptoms in postpartum women is of late significantly receiving growing scientific and clinical recognition.^6-8^

Of the general population of women giving birth worldwide, ∼6% are estimated to experience full childbirth-related PTSD (CB-PTSD).^7,8^ This translates to ∼8 million women affected in 2022. CB-PTSD can develop when the individual experiences an acute negative emotional and physiological response due to childbirth, and has been documented in both pre-term as well as full-term deliveries with healthy infant outcomes.^9-11^ Women at heightened risk are those with medically complicated deliveries, such as in cases of unscheduled/emergency Cesarean section,^12^ obstetrical complications,^6,13^ and maternal near-miss.^14,15^ Racial and ethnic disparities in experiences of childbirth trauma have also been documented;^16^ Black and Latinx women are nearly 3 times more likely to endorse acute stress response to childbirth.^16^ Altogether, ∼20% of high-risk individuals are likely to endorse CB-PTSD.^8,17^

CB-PTSD symptoms resemble the symptoms of general, non-postpartum PTSD. In accordance with the Diagnostic and Statistical Manual of Mental Disorders, Fifth Edition, (DSM-5), CB-PTSD entails trauma (childbirth)-specific intrusion and avoidance, negative alterations in cognitions and mood, and hyperarousal/hyper(-re)activity.^18^ Also, similarly to general PTSD, CB-PTSD can occur with symptoms of depression.^6^ However, unlike other forms of PTSD, CB-PTSD develops in temporal proximity to the birth of the child, and the child may become a traumatic reminder to his or her mother. A core complication of this maternal disorder is impairment in the development of mother-infant bonding. Maternal attachment problems are reported across the first postpartum year^19,20^ as well as reduced exclusive breastfeeding during an important time for child development.^11^ This suggests that CB-PTSD can impede early child development and result in significant public health costs.

An essential step in facilitating maternal psychological adjustment following traumatic deliveries relates to early and accurate identification of women with probable CB-PTSD. Accurate screening could be the first step in optimizing opportunities for effective interventions and allocating appropriate resources to targeted, at-risk individuals.^21^ The challenges of postpartum mental health screening involve, in part, women’s tendency to under-report their symptoms.^22^ Concerns of shame, stigma, and forced separation from infants, as well as poor awareness, hinder screening.^23-25^ This suggests that assessment of CB-PTSD derived primarily from patients’ reporting of symptoms could be limited by minimal introspective ability and desirability bias in reporting.

There is research interest that natural language derived from spontaneous word usage could serve as a marker of well-being and psychopathology.^26-28^ In the context of trauma, the personal memory of the event is a central contributor to PTSD etiology and maintenance.^29,30^ Accordingly, studies have demonstrated that the way in which individuals recall and narrate traumatic events, including the narrative language, relates to their post-traumatic stress symptom expression.^28,31-33^ This suggests that the words describing individuals’ narrative of the traumatic event, which represent the subjective, less filtered experience of the trauma, could represent post-trauma adjustment, even before extensive psychological processing and meaning making has occurred.^33^ Exciting developments in natural language processing (NLP) computational methods reveal that algorithms can analyze human language and extract insights similarly to how humans understand it, and in combination with machine learning (ML) models, they could be promising for informing the classification of psychiatric conditions.^34-36^ Recent transformer-based NLP methods^37^ enable algorithms to achieve state-of-the art results in understanding the contextual nuances of the language in written texts based on large quantities of natural text.^38^ NLP extracts and represents unstructured textual data as structured data that could then be used for generating ML classification models.^38^

Despite the potential of analyzing the rich text in personal narratives for examining the experience of childbirth, minimal quantitative investigations of word usage in birth narratives as markers of maternal mental health have been conducted.^18,39^ To our knowledge, research examining the utility of childbirth narratives combined with advanced text-based computational methods and ML to inform the identification of maternal mental health traumatic stress outcomes is lacking.

In this study, we collected short, unstructured narrative accounts of the personal and recent childbirth experiences from a total of 1,127 postpartum women. Using an NLP transformer-based algorithm and a developed ML classifier, we examined whether the text of narratives, alone, could be used to identify postpartum women with provisional CB-PTSD.

## Materials and Methods

### Study design

This investigation is part of a research study concerning the childbirth experience and maternal psychological sequelae during the COVID-19 era.^9^ Women who gave birth to a live baby in the last six months, and were at least 18 years old were enrolled, and they provided information about their mental health and childbirth experience via an anonymous web survey. Recruitment was during the period of 04/02/2020 to 12/29/2020, and was done using hospital announcements, social media, and professional organizations. The project received exemption from the Partners Healthcare (Massachusetts General Brigham) Human Research Committee (PHRC).

The sample in this study consists of 1,127 women who provided birth narratives, 1,111 of whom completed a PTSD symptom screen; of these, 995 (88.29%) provided written childbirth narratives of length 30+ words. On average, their maternal age was 32 ± 4.43 years and gestational period was 38.98 ± 1.71 weeks. 53.1% of participants were primiparas, and 69.6% gave birth via vaginal delivery. Around 10% (n = 86) had a positive PTSD Screen (PTSD Checklist for DSM-5, PCL-5 ≥ 31). Table 1 presents demographics and childbirth information for women with and without a positive screen.

**Table 1.** Demographics and childbirth factors by childbirth-related posttraumatic stress disorder status

| | CB-PTSD<br>(n=86) | | No CB-PTSD<br>(n=909) | | $\chi^2$ | OR (95% CI) |
| --- | --- | --- | --- | --- | --- | --- |
|  | % | n | % | n |  |  |
| **Maternal age** |  |  |  |  |  |  |
| <35 years | 67.4 | 58 | 68.2 | 620 | 0.02 | 0.97 (0.60-1.55) |
| **Education** |  |  |  |  |  |  |
| Less than bachelor's degree | 24.4 | 21 | 14.4 | 131 | 6.08* | 1.92 (1.13-3.25) |
| **Marital status** |  |  |  |  |  |  |
| Not married or domestic partnership | 10.5 | 9 | 5.9 | 54 | 2.71 | 1.85 (0.88-3.89) |
| **Household income** |  |  |  |  |  |  |
| <\$100,000 | 49.4 | 42 | 39.3 | 354 | 3.31 | 1.51 (0.97-2.36) |
| **Race and ethnicity** |  |  |  |  |  |  |
| Black and/or Hispanic or Latino | 15.3 | 13 | 7.4 | 67 | 6.45* | 2.25 (1.19-4.27) |
| **Prior mental health** | 52.3 | 45 | 32.2 | 293 | 14.14*** | 2.31 (1.48-3.60) |
| **Primiparity** | 66.3 | 57 | 51.8 | 471 | 6.60** | 1.83 (1.15-2.91) |
| **Pain medication** | 82.6 | 71 | 76.5 | 695 | 1.61 | 1.45 (0.81-2.59) |
| **Medication induction** | 57.0 | 49 | 45.8 | 416 | 3.93* | 1.57 (1.00-2.45) |
| **Obstetrical complications** | 54.7 | 47 | 24.1 | 219 | 37.46*** | 3.80 (2.42-5.96) |
| **Sleep deprivation** | 84.2 | 64 | 65.5 | 539 | 11.04*** | 2.81 (1.49-5.29) |
| **Mode of delivery** |  |  |  |  |  |  |
| Vaginal | 45.3 | 39 | 71.9 | 654 | 26.29*** | 0.32 (0.21-0.51) |
| Planned Cesarean | 8.1 | 7 | 12.9 | 114 | 1.43 | 0.62 (0.28-1.37) |
| Unplanned Cesarean | 46.5 | 40 | 15.5 | 141 | 50.74*** | 4.74 (2.99-7.50) |
| **Premature delivery** | 14.0 | 12 | 5.7 | 52 | 8.85** | 2.67 (1.37-5.23) |
| **Sense of danger for self or newborn's life** | 53.5 | 46 | 14.5 | 131 | 81.48*** | 6.80 (4.28-10.79) |
| **Skin-to-skin** | 65.1 | 56 | 90.7 | 824 | 50.84*** | 0.19 (0.12-0.31) |
| **Rooming in** | 72.1 | 62 | 92.8 | 831 | 41.40*** | 0.20 (0.12-0.34) |
| **NICU admission** | 30.2 | 26 | 82.7 | 96 | 27.96*** | 3.65 (2.20-6.05) |
| **Acute stress in childbirth** | 79.1 | 68 | 17.4 | 158 | 170.32*** | 17.96 (10.39-31.03) |
|  | *M* | *SD* | *M* | *SD* |  |  |
| **Mean postpartum study completion (months)** | 3.13 | 1.96 | 2.45 | 1.75 |  |  |

Note for Table 1. CB-PTSD = provisional childbirth-related posttraumatic stress disorder (PCL-5 $\geq 31$ ); Sleep deprivation = $<6$ hours of sleep the night before birth; Premature delivery = $<37$ weeks gestation age; NICU = neonatal intensive care unit; Sense of danger refers to during/immediately after childbirth, at least moderate degree (PCL-5 A1); Acute stress refers to clinically significant immediate emotional and psychological response to personal childbirth (PDI $\geq 17$ ).
OR = Odd ratios, 95% CI = 95% confidence interval.
\* $p < .05$ , \*\* $p < .01$ , \*\*\* $p < .001$ .
Differences in sample sizes are due to missing data.

### Measures

*Narratives of childbirth* were collected as written (typed) unstructured, open-ended accounts of each participant’s personal and recent childbirth experience. Narratives were collected in a free recall paradigm in which participants were instructed as follows: “Please provide a brief description of your recent childbirth experience in your own words, with a focus on the most distressing aspects of your experience, if applicable.”

*PTSD symptoms specific to childbirth* were measured using the PTSD Checklist for DSM-5 (PCL-5).^40^ This is the standard self-report measure to assess the presence and severity of the 20 DSM-5 PTSD symptoms following an index traumatic event endorsed over the past month using a 0 (not at all) to 4 (extremely) scale. Participants were instructed to report on their symptoms in regard to recent childbirth. The PCL-5 has strong correspondence with clinician diagnostic assessments and is used to determine provisional PTSD diagnosis,^41^ with a reported clinical cutoff of 31.^42^ Reliability of the measure was high (Cronbach’s alpha = 0.91).

### Modeling methods

To analyze narrative text, we represented sentences (narratives) as dense, low-dimensional vectors termed ‘embeddings’, using the `all-mpnet-base-v2` pre-trained Sentence-Transformers NLP model.^43^ This model maps sentences and paragraphs to a fixed size of 768-dimensional dense vector. It fine-tuned Microsoft’s pretrained `mpnet-base` NLP model^44^ on a dataset of 1 billion sentence pairs to identify sentence similarity (essential to our developed method), and it was reported to produce the highest average performance^45^ on encoding sentences over 14 NLP tasks, compared with other NLP models.

We developed an ML model that utilizes the output (sentence embedding vectors) of the `all-mpnet-base-v2` NLP model to identify provisional CB-PTSD via narrative classification. The developed ML model was trained to classify childbirth narratives as markers of endorsement, or no endorsement, of CB-PTSD. Appendix A presents the 4 steps to build and test our model, and Appendix B provides sensitivity analysis of our model.

## Results

### Descriptive

Following the data processing (Steps 1 and 2 in Appendix A), for Class 1 (CB-PTSD) and Class 0 (No CB-PTSD), the mean and median word count (WC) were 191.91 and 142, and 154.61 and 106, respectively. A t-test analysis revealed that participants of the CB-PTSD class used more words to depict their childbirth experiences than those of the No CB-PTSD class (*t* = 2.30, *df* = 111.99, *p* = 0.02) (Figure 1).

**Figure 1.**
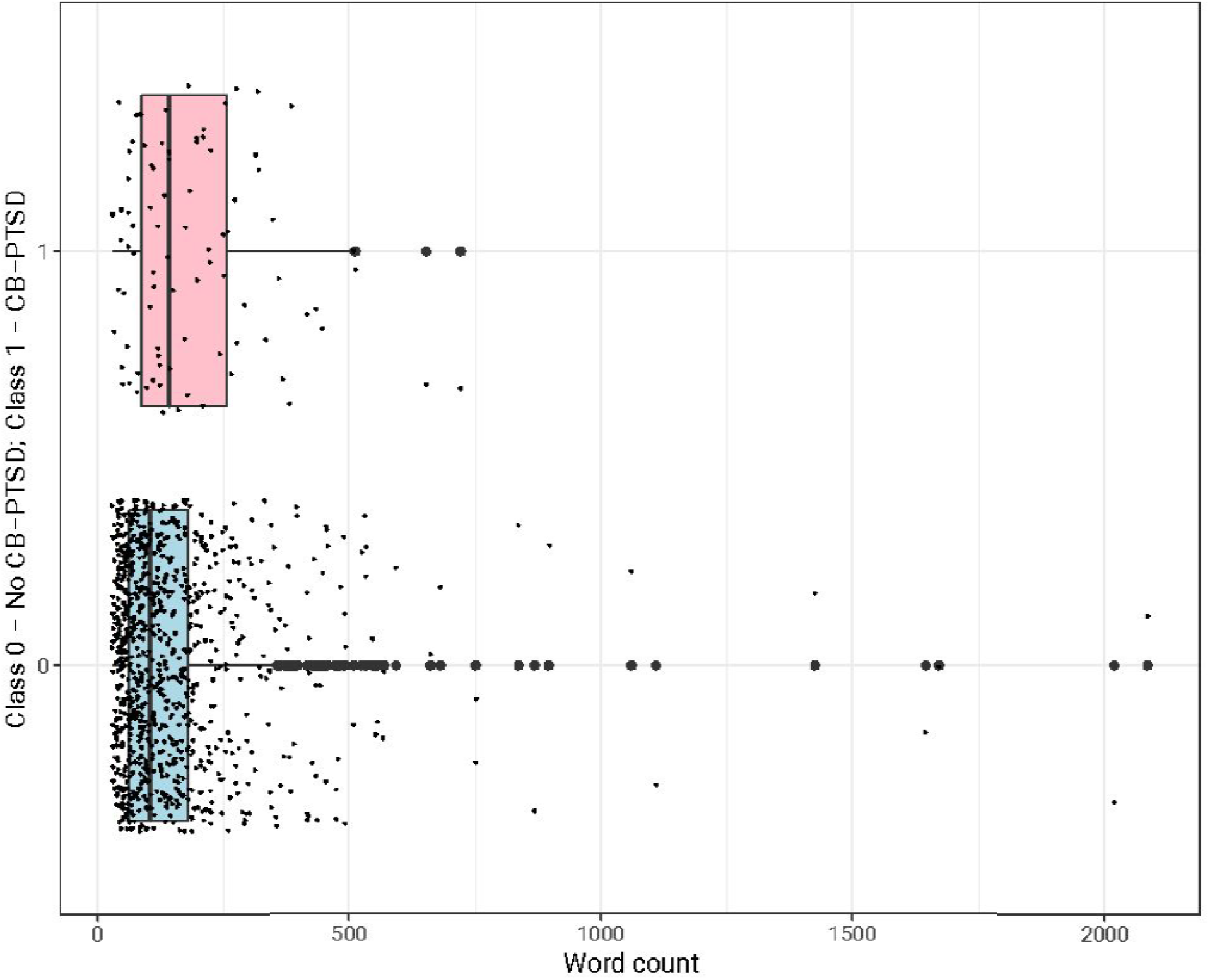
Number of words in childbirth narratives by childbirth-related PTSD status. Boxplots display word count in narratives for CB-PTSD (Class 1, PCL-5 ≥ 31, pink) and No CB-PTSD (Class 0, PCL-5 < 31, light blue). Dots are data points (narratives’ word counts) shifted by a random value. The mean word count (WC) for Class 1 is 191.91, and for Class 0 is 142. The median WC for Class 1 is 154.61, and for Class 0 is 106. A t-test revealed that participants of Class 1 used more words to depict their birth narrative than those of Class 0 (*t* = 2.30, *df* = 111.99, *p* = 0.02).

### Machine learning modeling analysis

We labeled narratives associated with PCL-5 ≥ 31 as “CB-PTSD” (Class 1), and PCL-5 < 31 as “No CB-PTSD” (Class 0) (Step 1, Appendix A). Next, we discarded short narratives (<30 words) to allow meaningful learning of word patterns,^46^ resulting in the removal of 116 narratives (Step 2.1, Appendix A). Then, we balanced the dataset using down-sampling by randomly sampling the majority Class 0 to fit the size of the minority Class 1, resulting in 86 narratives in each class. We constructed the train and test datasets as described in Step 2.2 in Appendix A. We repeated this step (and the following steps) 10 times to capture different narratives for creating an accurate representation of Class 0 and Class 1.

Next, we created three sets of sentence pairs using the train set. Set #1: all possible pairs of sentences (2,145) in Class 1 (CB-PTSD); Set #2: all possible pairs of sentences (2,145) in Class 0; and Set #3: pairs of sentences (4,290), one randomly selected from Class 1 and another randomly selected from Class 0. We labeled Sets #1 and #2 as positive examples since they contained semantically similar pairs of sentences (either a pair of narratives of participants with or without CB-PTSD). We labeled Set #3 as negative examples since they contained pairs of non-semantically similar pairs of sentences. This data augmentation process produced a total of 8,580 training examples generated from the train set (Step 3.1, Appendix A).

We mapped each narrative using the `all-mpnet-base-v2` model into a 768-dimensional vector. Then, we standardized these vectors by removing the mean and scaling to unit variance. Finally, we computed the absolute element-wise difference between each of the 8,580 embedding vectors of pair of sentences *u, v* in Sets #1 to #3 of the train set (Step 3.1, Appendix A), such that *z* = (|*emb*(*u*) − *emb*(*v*)|) (Step 3.2, Appendix A).

Using the 8,580 calculated *z* vectors, we trained a DFNN model to classify pairs of sentences in Sets #1 to #3 as semantically similar or not (Step 3.3, Appendix A). For training, we used the Keras Python library and constructed a DFNN with an input layer of 768 neurons, two hidden layers of 400 and 50 neurons, and an output neuron. All layers had a ReLU activation function, except for the output neuron with a Sigmoid activation function. We used 150 epochs, applying the Adam optimizer with a learning rate of 3e^-5^, a batch size of 64, and binary cross-entropy loss to monitor training performance. To avoid overfitting, we stopped training when there was no loss improvement for three consecutive epochs. We used 20% of the train dataset for validation during the training process.

Finally, we tested and compared the performance of the developed model against a baseline model using a 10-fold CV (Step 4, Appendix A). As a baseline model, we fine-tuned the `all-mpnet-base-v2` NLP model using the train dataset on a downstream task of classifying narratives into Class 0 or Class 1. We used the Sentence-Transformers Python library within the HuggingFace Hub with the following parameters: learning rate=2e^-5^, batch size=16, epochs=50, weight decay=0.001.

The results of applying the baseline model to the test set and our model are presented in Table 2.

**Table 2.** Comparison of the average (10 different seeds) performance classification results of the developed model vs. a baseline model. Both models use exclusively text features to identify childbirth-related PTSD.

| **Model** | **AUC**<br>**\*** | **F1-score\*\*** | **Sensitivity** | **Specificity** |
| --- | --- | --- | --- | --- |
| Baseline model | 0.53 | 0.52 | 0.66 | 0.41 |
| The developed model | 0.75 | 0.76 | 0.80 | 0.70 |

\*A model with an AUC of 0 suggests no ability to diagnose patients, and 1 a perfect diagnosis.
\*\* F1-score ranges between 0 to 1 (a perfect classification).

The results of the baseline model emphasize the problem of training an ML classifier with a small number of examples. In contrast, our model was able to overcome this problem by using 8,580 training examples, outperforming the baseline model. Our model for provisional CB-PTSD classification derived from birth narratives achieved overall good performance.

### Word category analysis

To examine the use of specific word categories in the birth narratives and their potential relation to CB-PTSD status, we used the Linguistic Inquiry and Word Count (LIWC) software,^47^ which uses different validated dimensions to classify words into categories. It compares a word from the natural text input to a dictionary of pre-defined words, and classifies the identified word into a predefined dimension.^47^

We examined the frequency of specific word categories in childbirth narratives that were previously shown to represent trauma narratives of individuals with PTSD.^32,48^ Using LIWC, we examined the frequency of: ‘Affect’, ‘Anger’, ‘Anx’, ‘Bio’, ‘Body’, ‘Cause’, ‘Cogproc’, ‘Death’, ‘Feel’, ‘Filler’, ‘Health’, ‘Hear’, ‘I’, ‘Insight’, ‘Negemo’, ‘Percept’, ‘Posemo’, ‘Pronoun’, ‘Sad’, ‘See’, ‘We’, and ‘You’. A Wilcoxon test for differences in the word frequency revealed that participants of the CB-PTSD class used fewer positive emotions, and more negative emotions as well as body- and death-related words in their birth narratives, compared with the No CB-PTSD class (Figure 2).

**Figure 2.**
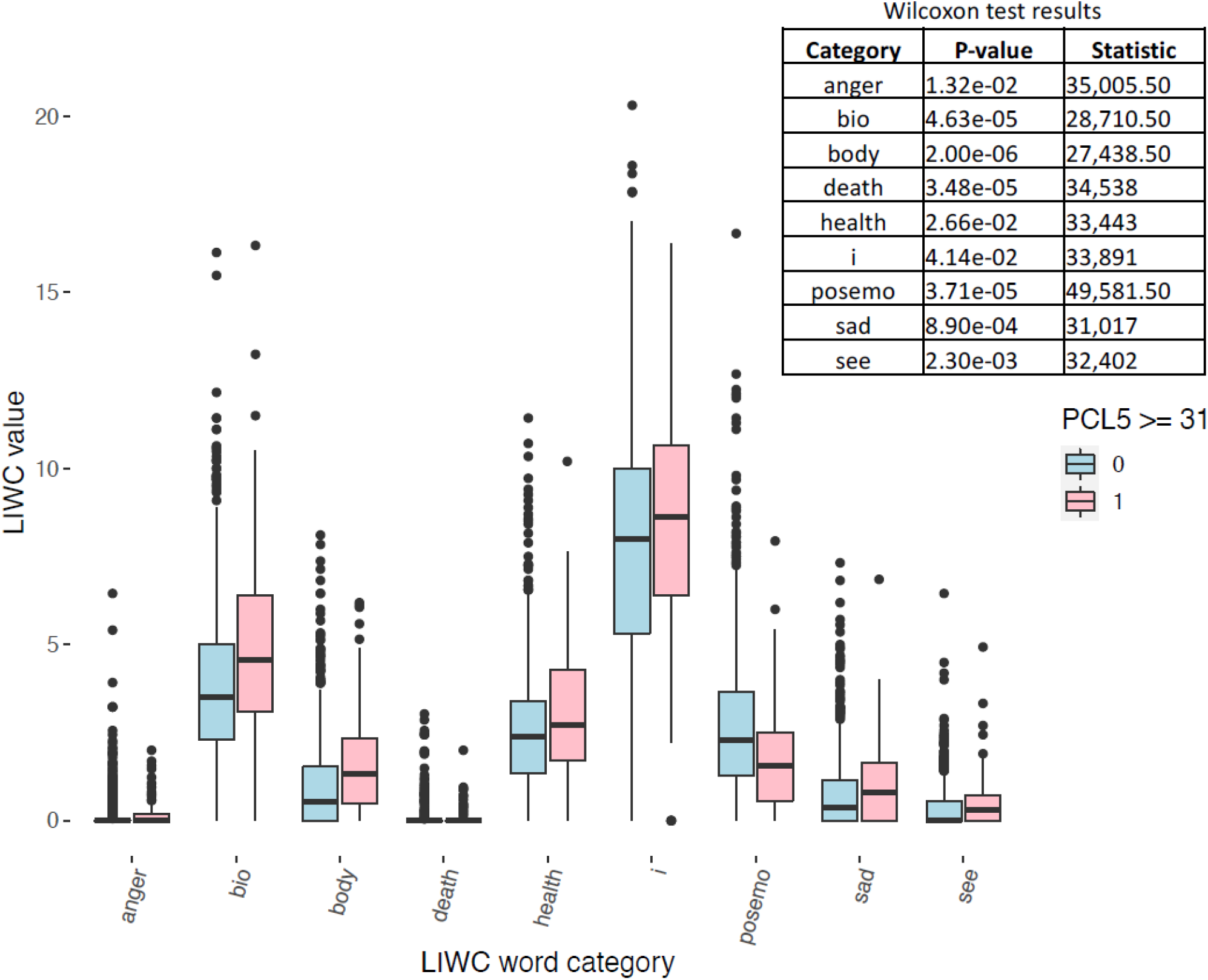
Frequency of words in childbirth narratives by childbirth-related PTSD status. Distribution of word frequencies (LIWC value) per CB-PTSD Class (Class 1, CB-PTSD, pink; and Class 0, No CB-PTSD, light blue). PTSD was measured by PTSD Checklist for DSM-5 (PCl-5 ≥ 31). The table in the figure elaborates significant results of a Wilcoxon rank sum test with continuity correction between a word category in Class 0 and Class 1. X-axis label ‘i’ is the first-person pronoun “I”.

### Comment

#### Principal findings

This study shows that advanced machine learning (ML) methods for analyzing free text have the potential to identify women with provisional CB-PTSD following childbirth based on short, unstructured personal childbirth written narrative accounts. This simple data collection method appears feasible and efficient for collecting information from a large number of postpartum women during a sensitive time period, and may overcome the inherent barriers of relying on medical record data to identify at-risk women.^49-51^ In our final model, up to 80% (sensitivity) of women who likely meet provisional CB-PTSD criteria could be accurately identified based on word usage in their narratives, and 70% (specificity) of those not endorsing the condition could be identified as such.

### Results in the context of what is known

To our knowledge, this is the first study to use childbirth narratives accounts and state-of-the-art NLP algorithms combined with ML models for the identification via classification of a maternal mental health condition in general.^52^ Research using ML models for the classification of CB-PTSD is largely lacking. Only a few studies have tested the utility of ML models for CB-PTSD identification.^53,54^ While our model’s performance is comparable to the reported models, those models are informed by relatively extensive data such as information derived from medical records^53^ and/or structured questionnaires.^53,54^ In contrast, the use of freely generated childbirth narratives may not only have the advantage of being a more accessible data collection method, but also entails self-disclosure and narrative construction, which both have positive implications in the processing of traumatic events and facilitating psychological adaptation.^55-58^

### Clinical implications

Although early and mass screening for CB-PTSD would likely improve diagnosis rates and facilitation of treatment, there are no recommended medical protocols for CB-PTSD screening in hospitals and health clinics. The opportunity to screen women when they are still in contact with obstetrical providers is important, as such contact appears much more difficult to establish later, when disorders can become chronic and often comorbid,^6^ and hence, more difficult to treat.^59,60^ Establishing the potential accuracy of non-invasive and low-cost data collection based on the words in childbirth narratives for the identification of women with provisional CB-PTSD could serve as an important first step to ultimately complement more extensive clinical assessments and other biologically-oriented methods.^10^ Collection of short written childbirth accounts could be done remotely, with the possibility of introducing an assessment with minimal burden during an acute period of rapid physiological and psychological adjustment. It may have the potential to be readily implemented in routine obstetrical care.

### Research implications

We applied an advanced NLP model for analyzing the text in personal childbirth narratives. Although NLP has been increasingly used to analyze free-text medical data, the application of NLP to birth narratives for identification of CB-PTSD has not previously been done. We report an AUC of 0.75 that accords with other studies of non-postpartum psychiatric classifications that used NLP models.^61^ The uniqueness of the method applied here is the development of a pairwise sentence similarity classification model that uses a Sentence-Transformers NLP model to embed narratives, and uses these embeddings to train a neural network classifier for CB-PTSD identification. Our results suggest that computational models that can understand the context of the language, as humans do, are promising for guiding the detection of maternal psychiatric morbidity from narrative birth-related text. Future studies are needed to replicate our results.

### Strengths and limitations

This study presents the first use of NLP to identify a maternal mental health condition from women’s personal written accounts of their childbirth experience collected remotely. Our model demonstrated good performance; for future work, combining such narratives with information in patients’ medical records and physiological responses to childbirth^10^ may enhance the accuracy in forecasting the maternal mental health outcome following traumatic experiences of childbirth..

Several limitations to this work are worth noting. Although we used the well-validated PTSD checklist for DSM-5 that corresponds strongly with diagnostic measures^62^ to determine provisional CB-PTSD status, clinician assessments were not performed. The samples largely represent middle-class American women, and this warrants replication of the work in more diverse populations. The model was developed and tested on the collected dataset; however, we used a pre-trained NLP model to represent narratives as vectors, and this may not fully capture the language nuances in childbirth narratives; this necessitates future work in training a new NLP model on a larger narrative dataset.

## Conclusions

In summary, this study is a proof of concept of the potential utility of the text in childbirth narratives, alone, to inform the detection of early signs of maternal PTSD following childbirth using state-of-the-art NLP, and ML models. As psychiatric morbidity in the transition to motherhood remains a public health concern,^63,64^ more research is needed to guide the development of tools for the accurate and early screening of women likely to endorse a mental illness following childbirth.

## Data Availability

All data produced in the present study are available upon reasonable request to the authors.

## Disclosure Statement

The authors report no conflict of interest.

## Funding

Dr. Sharon Dekel was supported by a grant from the National Institute of Child Health and Human Development (R21HD100817) and an ISF award from the Massachusetts General Hospital Executive Committee on Research. Dr. Kathleen Jagodnik was supported by the Mortimer B. Zuckerman STEM Leadership Postdoctoral Fellowship Program. The sponsors were not involved in study design; in the collection, analysis or interpretation of data; in the writing of the report; or in the decision to submit this article for publication.

## Acknowledgment

The authors would like to thank Prof. James W. Pennebaker for his generous contribution in performing the Linguistic Inquire Word Count (LIWC) analysis. Dr. Pennebaker is without conflict and agreed to be acknowledged.

## Notes

### Competing Interest Statement

The authors have declared no competing interest.

### Author Declarations

The project received exemption from the Partners Healthcare (Massachusetts General Brigham) Human Research Committee (PHRC).

## References

1. Sommerlad S, Schermelleh-Engel K, La Rosa VL, Louwen F, Oddo-Sommerfeld S. Trait anxiety and unplanned delivery mode enhance the risk for childbirth-related post-traumatic stress disorder symptoms in women with and without risk of preterm birth: A multi sample path analysis. Plos One 2021;16(8):e0256681.

2. Türkmen H, Yalniz Dilcen H, Akin B. The Effect of labor comfort on traumatic childbirth perception, post-traumatic stress disorder, and breastfeeding. Breastfeeding Medicine 2020; 15(12):779–788.

3. Boorman RJ, Devilly GJ, Gamble J, Creedy DK, Fenwick J. Childbirth and criteria for traumatic events. Midwifery 2014;30(2):255–261.

4. Ayers S. Delivery as a traumatic event: prevalence, risk factors, and treatment for postnatal posttraumatic stress disorder. Clin Obstet Gynecol 2004;47(3):552–567.

5. Dekel S, Gilberston M, Orr S, Rauch S, Nellie W, Pitman R. Trauma and posttraumatic stress disorder. In T.A. Stern, M. Fava, T. Wilens, J.F. Rosenbaum (Eds.): Mass General Hosp Compreh Clin Psych 2nd Ed., Philadelphia, PA, Elsevier, 2016.

6. Dekel S, Ein-Dor T, Dishy GA, Mayopoulos PA. Beyond postpartum depression: posttraumatic stress-depressive response following childbirth. Arch Wom Ment Health 2020;23(4):557–564.

7. Dekel S, Stuebe C, Dishy G. Childbirth induced posttraumatic stress syndrome: a systematic review of prevalence and risk factors. Front Psychol 2017;8:560.

8. Yildiz PD, Ayers S, Phillips L. The prevalence of posttraumatic stress disorder in pregnancy and after birth: A systematic review and meta-analysis. J Affect Disord 2017;208:634–645.

9. Mayopoulos GA, Ein-Dor T, Dishy GA et al. COVID-19 is associated with traumatic childbirth and subsequent mother-infant bonding problems. J Affect Disord 2021;282: 122–125.

10. Chan SJ, Thiel F, Kaimal AJ, Pitman RK, Orr SP, Dekel S. Validation of childbirth-related posttraumatic stress disorder using psychophysiological assessment. Am J of Obstet Gynecol 2022:S0002-9378.

11. Chan SJ, Ein-Dor T, Mayopoulos P et al. Risk factors for developing posttraumatic stress disorder following childbirth. Psychiatry Res 2020;290:113090.

12. Dekel S, Ein-Dor T, Berman Z, Barsoumian IS, Agarwal S, Pitman RK. Delivery mode is associated with maternal mental health following childbirth. Arch Wom Ment Health 2019;22(6):817–824.

13. Kazato M. Screening for childbirth-related posttraumatic stress disorder using the City Birth Trauma Scale: A pilot project (2022). Doctor of Nursing Practice Final Manuscripts. 187.

14. England N, Madill J, Metcalfe A et al. Monitoring maternal near miss/severe maternal morbidity: a systematic review of global practices. PloS One 2020;15(5):e0233697.

15. Soma-Pillay P, Anthony J, Mandondo SD. Cardiac disease in pregnancy: when to raise the ‘red flag’. South Afr Med J 2018;108(11):901–906.

16. Iyengar AS, Ein-Dor T, Zhang EX, Chan SJ, Kaimal AJ, Dekel S. Increased traumatic childbirth and postpartum depression and lack of exclusive breastfeeding in Black and Latinx individuals. Int J Gynecol Obstetr 2022.

17. Polachek IS, Dulitzky M, Margolis-Dorfman L, Simchen MJ. A simple model for prediction postpartum PTSD in high-risk pregnancies. Arch Wom Ment Health 2016;19(3):483–490.

18. Thiel F, Berman Z, Dishy GA et al. Traumatic memories of childbirth relate to maternal postpartum posttraumatic stress disorder. J And Discord 2021;77:102342.

19. Dekel S, Thiel F, Dishy G, Ashenfarb AL. Is childbirth-induced PTSD associated with low maternal attachment?. Arch Women’s Mental Health 2019;22(1):119–122.

20. Kjerulff KH, Attanasio LB, Sznajder KK, Brubaker LH. A prospective cohort study of post-traumatic stress disorder and maternal-infant bonding after first childbirth. J Psychosom Res 2021;144110424.

21. Berman Z, Thiel F, Kaimal AJ, Dekel S. Association of sexual assault history with traumatic childbirth and subsequent PTSD. Arch Wom Ment Health 2021;24(5):767–771.

22. Anokye R, Acheampong E, Budu-Ainooson A, Obeng EI, Akwasi AG. Prevalence of postpartum depression and interventions utilized for its management. Ann Gen Psych 2018;17(1):1–8.

23. Burval J, Kerns R, Reed K. Treating postpartum depression with brexanolone. Nursing 2022, 2020;50(5):48–53.

24. Jones A. Postpartum help-seeking: the role of stigma and mental health literacy. Matern Child Health J 2022;26(5):1030–1037.

25. Lackie ME, Parrilla JS, Lavery BM et al. Digital health needs of women with postpartum depression: focus group study. J Med Internet Res 2021;23(1):p. e18934.

26. Boyd RL. The multifaceted measurement of the individual through language (Doctoral dissertation), The University of Texas at Austin (2017).

27. Wardecker BM. Correlates and Predictors of Emotion Language and Well-Being in Stressful and Traumatic Contexts (Doctoral dissertation), University of Michigan, Horace H. Rackham School of Graduate Studies (2016).

28. Pennebaker JW, Mehl MR, Niederhoffer KG. Psychological aspects of natural language use: our words, our selves. Annu Rev Psychol 2003;54:547–577.

29. Engelhard IM, McNally RJ, van Schie K. Retrieving and modifying traumatic memories: recent research relevant to three controversies. Curr Dir Psychol Sci 2019;28(1):91–96.

30. Brewin CR. Memory and forgetting. Curr Psych Rep 2018;20(10):1–8.

31. Crespo M, Fernández-Lansac V. Memory and narrative of traumatic events: a literature review. Psych Trauma: Theory, Res, Pract, Pol 2016;8(2):149.

32. Dekel S, Bonanno G. Changes in trauma memory and patterns of posttraumatic stress. Psych Trauma: Theory, Res, Pract, Pol 2013;5(1):26–34.

33. O’Kearney R, Perrott K. Trauma narratives in posttraumatic stress disorder: a review. J Trauma Stress 2006;19(1):81–93.

34. Calvo RA, Milne DN, Hussain MS, Christensen H. Natural language processing in mental health applications using non-clinical texts. Nat Lang Eng 2017;23(5):649–685.

35. He Q, Veldkamp BP, Glas CA, de Vries T. Automated assessment of patients’ self-narratives for posttraumatic stress disorder screening using natural language processing and text mining. Assessment 2017;24(2):157–172.

36. Tsui FR, Shi L, Ruiz V et al. Natural language processing and machine learning of electronic health records for prediction of first-time suicide attempts. JAMIA Open 2021;4(1):ooab011.

37. Devlin J, Chang MW, Lee K, Toutanova K. Bert: Pre-training of deep bidirectional transformers for language understanding. arXiv preprint 2018:1810.04805

38. M.C. Monuteaux, C. Stamoulis, C. Machine Learning: A Primer for Child Psychiatrists. Journal of the American Academy of Child and Adolescent Psychiatry 2016;55(10):835–836.

39. Santoro E, Stagni-Brenca E, Olivari MG, Confalonieri E, Di Blasio P. Childbirth narratives of women with posttraumatic stress symptoms in the postpartum period. J Obstet Gyn & Neonatal Nursing 2018;47(3):333–341.

40. Weathers FW, Litz BT, Keane TM, Palmieri PA, Marx BP, Schnurr PP. The PTSD checklist for DSM-5 (PCL-5) scale. National Center for PTSD (2013) Available at http://www.ptsd.va.gov, Accessed 6th August 2022

41. Wortmann JH, Jordan AH, Weathers FW et al. Psychometric analysis of the PTSD Checklist-5 (PCL-5) among treatment-seeking military service members. Psychology Assessment 2016;28(11):1392.

42. Krüger-Gottschalk A, Knaevelsrud C, Rau H et al. The German version of the Posttraumatic Stress Disorder Checklist for DSM-5 (PCL-5): psychometric properties and diagnostic utility. BMC Psychiatry 2017;17:379.

43. Hugging Face 2022. all-mpnet-base-v2. https://huggingface.co/sentence-transformers/all-mpnet-base-v2. Accessed 8/11/2022.

44. Song K, Tan X, Qin T, Lu J, Liu TY. Mpnet: masked and permuted pre-training for language understanding. Adv Neural Info Proc Syst 2020;33:16857–16867.

45. Sbert 2022. Model Comparisons Table. https://www.sbert.net/_static/html/models_en_sentence_embeddings.html. Accessed 8/11/2022.

46. Bartal A, Lachmann A, Clarke DJB, Seiden AH, Jagodnik KM, Ma’ayan A. EnrichrBot: Twitter bot tracking tweets about human genes. Bioinformatics 2020;36(12):3932–3934.

47. Pennebaker JW, Boyd RL, Jordan K, Blackburn K. The development and psychometric properties of LIWC2015. Austin, TX: University of Texas at Austin (2015).

48. Chin SW, Chong ST, Musa ABB, Loh KS. The data management of the language of trauma narrative communication. In 2021 IEEE Global Eng Educat Conf (EDUCON) (2021):1502–1508.

49. Hammack-Aviran CM, Brelsford KM, McKenna KC, Graham RD, Lampron ZM, Beskow LM. Research use of electronic health records: patients’ views on alternative approaches to permission. AJOB Emp Bioeth 2020;11(3):172–186.

50. Ta CN, Weng C. Detecting systemic data quality issues in electronic health records. Studies in health technology and informatics 2019;264:383.

51. Cowie MR, Blomster JI, Curtis LH et al. Electronic health records to facilitate clinical research. Clinical Research in Cardiology 2017;106(1):1–9.

52. Saqib K, Khan AF, Butt ZA. Machine learning methods for predicting postpartum depression: scoping review. JMIR Mental Health 2021;8(11):e29838.

53. Orovas C, Orovou E, Dagla M et al. Neural Networks for Early Diagnosis of Postpartum PTSD in Women after Cesarean Section. Appl Sci 2022;12(15):7492.

54. Rousseau S, Polachek IS, Frenkel TI. A machine learning approach to identifying pregnant women’s risk for persistent post-traumatic stress following childbirth. J Affect Disord 2022;296:136–149.

55. Zheng L, Lu Q, Gan Y. Effects of expressive writing and use of cognitive words on meaning making and post-traumatic growth. J Pac Rim Psychol 2019; 13:e5.

56. Sloan DM. Self-disclosure and psychological well-being. In Maddux JE & Tangney JP (Eds.), Social psychological foundations of clinical psychology 2010:212–225. The Guilford Press.

57. Baikie KA, Wilhelm K. Emotional and physical health benefits of expressive writing. Adv Psychiatr Treat 2005;11(5):338–346.

58. Pennebaker JW, Seagal JD. Forming a story: The health benefits of narrative. J Clin Psychol 1999;55(10):1243–1254.

59. Bengel J, Becker-Nehring K, Hillebrecht J. Early psychological interventions. In Trauma Sequelae (2022): 175–202. Springer, Berlin, Heidelberg.

60. Roberts NP, Kitchiner NJ, Kenardy J, Lewis CE, Bisson JI. Early psychological intervention following recent trauma: a systematic review and meta-analysis. Eur J Psychotraum 2019;10(1):1695486.

61. Cohen J, Wright-Berryman J, Rohlfs L et al. A feasibility study using a machine learning suicide risk prediction model based on open-ended interview language in adolescent therapy sessions. Int J Env Res Public Health 2020;17(21):8187.

62. Bovin MJ, Marx BP, Weathers FW et al. Psychometric properties of the PTSD checklist for Diagnostic and Statistical Manual of Mental Disorders – Fifth Edition (PCL-5) in veterans. Psych Assess 2016;28:1379–1391.

63. Bailey L, Gaskin K. Spotlight on maternal mental health: a prepandemic and postpandemic priority. Evid-Based Nurs 2021;24(2):29–30.

64. Howard LM, Khalifeh H. Perinatal mental health: a review of progress and challenges. World Psychiatry 2020;19(3):313–327.

65. Reimers N, Gurevych I. Sentence-bert: sentence embeddings using siamese bert-networks. arXiv preprint 2019:1908.10084.

