## Appendices and Supplementary Figures for "Identifying Women with Post-Delivery Posttraumatic Stress Disorder using Natural Language Processing of Personal Childbirth Narratives"

==========================================================================================

### Appendix A: Steps to Build and Test the Developed Model

The following four steps describe how we built and tested the developed model (also summarized in Supplemental Figure 1a):

Step 1. *Define a PCL-5 cutoff score*.

We labeled each narrative as *Class 1*: probable CB-PTSD ('CB-PTSD') based on PCL-5 ≥ 31, or *Class 0*: no probable CB-PTSD ('No CB-PTSD') based on PCL-5 < 31 (see also Appendix B: Sensitivity Analysis section for additional tested PCL-5 cutoffs).

Step 2. *Data preparation*.

2.1 We discarded narratives with fewer than 30 words from the dataset. To handle imbalance in the analyzed dataset (due to small representation of cases with PCL-5 ≥ 31), we randomly sampled the majority Class 0 to fit the size of the minority Class 1.

2.2 Using the balanced dataset (Step 2.1), we randomly selected 75% of the narratives for training our model, and 25% for testing our model (see also Appendix B: Sensitivity Analysis section for additional assessed train/test splits).

Step 3. *Develop a machine learning classifier that utilizes NLP features.*

Using the train set, we analyzed pairwise narrative (sentence) similarity, following the Siamese networks approach[65] that learns to identify semantically similar pairs of sentences. This approach allowed us to generate multiple training examples since there are $\frac{n(n-1)}{2}$ possible combinations for $n$ sentences, thus addressing the challenge of training an ML model with a low number of examples, as in Class 1. The following three steps describe the model development.

3.1. Each pair of sentences in Class 1, and each pair of sentences in Class 0, was labeled as **positive examples**, indicating semantically *similar* sentences of individuals with (Set 1), or without (Set 2) CB-PTSD, respectively. Next, **negative examples** (Set 3) of the same size as the positive examples sets (|Set 1| + |Set 2|) were created by randomly selecting pairs of sentences, one from Class 1, and the other from Class 0, indicating semantically *non-similar* sentences.

3.2. Using the `all-mpnet-base-v2` model, each sentence in Class 1 and Class 0 was mapped into a dense vector space. Then, for each Set 1 to 3, we computed a vector $z$ of the absolute element-wise difference between the embedding (emb) vectors of each pair of sentences $(u, v)$, selected in Step 3.1, such that $z = (|emb(u) - emb(v)|)$.

3.3. We trained a densely connected feedforward neural network (DFNN) to classify pairs of sentences (by processing vector $z$) as semantically similar, or not (Supplemental Figure 1b).

Step 4. *Test model performance.*

We compared the performance of our model to a baseline model (see Results section) using 10-fold cross-validation (CV), and reported the area under the receiver operating characteristic curve (AUC), F1-score, Sensitivity, and Specificity performance measures on the test set. For testing our model on a newly unseen narrative $S$ in the test set, we first compute its embeddings using the `all-mpnet-base-v2` model. Next, we compute the average embedding vector ($\bar{v}_n$) of all train narratives in Class 0, and the average embedding vector ($\bar{v}_p$) of all train narratives in Class 1. To decide the class of $S$, we compute $z_n = (|emb(S) - \bar{v}_n|)$, and $z_p = (|emb(S) - \bar{v}_p|)$. Then, we apply our model to $z_n$ and $z_p$, and compare its output, i.e., compare the similarity likelihood of $emb(S)$ to $\bar{v}_p$ with the similarity likelihood of $emb(S)$ to $\bar{v}_n$. If $z_p > z_n$, we say that $S \in$ Class 1, else $S \in$ Class 0 (Supplemental Figure 2). Intuitively, our model should assign a larger similarity likelihood between an embedded narrative of an individual with CB-PTSD to vector $\bar{v}_p$ than to vector $\bar{v}_n$.

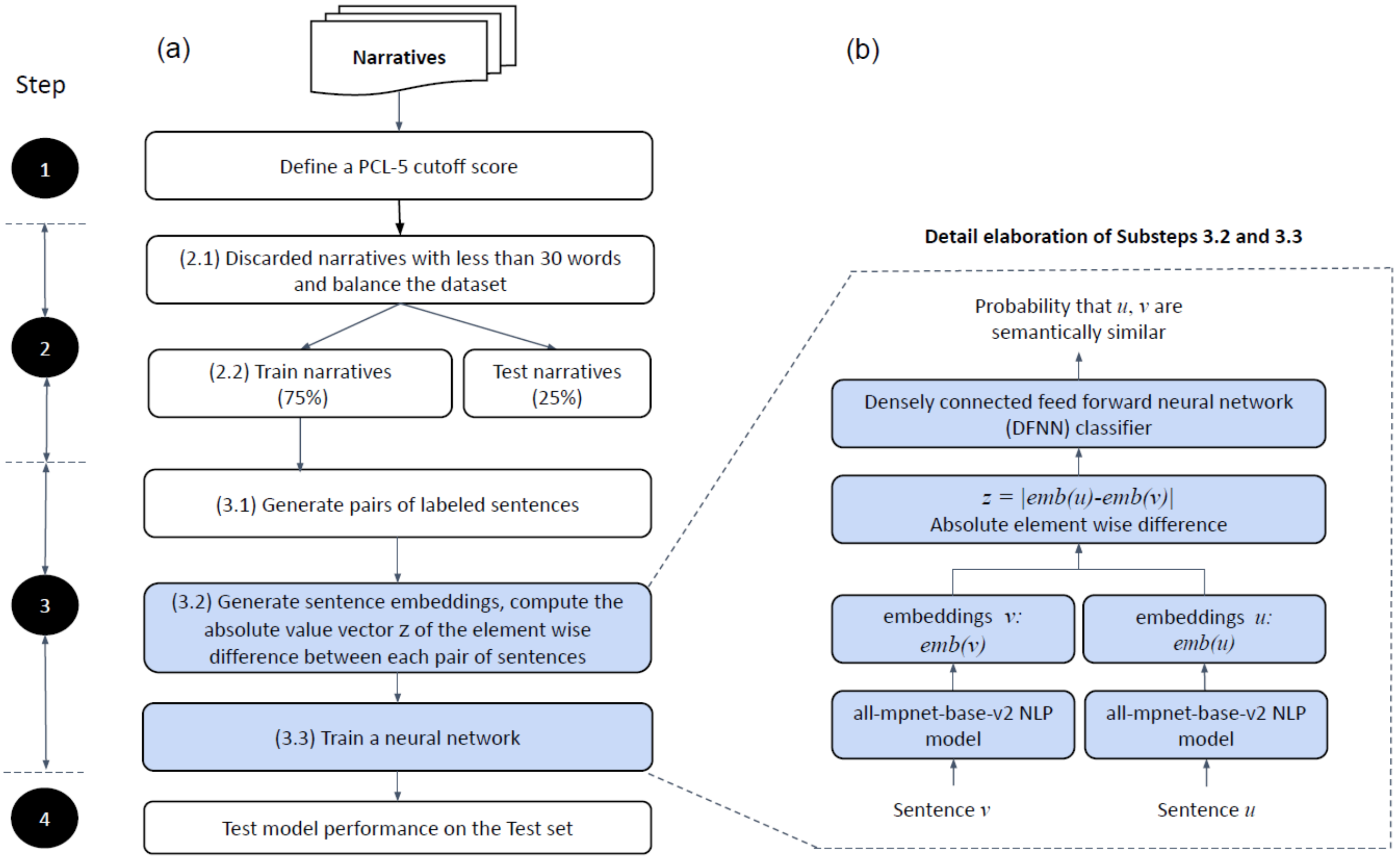


Supplemental Figure 1. Steps of the machine learning classifier model development for the identification of maternal childbirth-related post-traumatic stress disorder (CB-PTSD). (a) Left panel: An illustration summary of the four steps of the model. (b) Right panel: Training a neural network to identify pairwise sentence similarity.

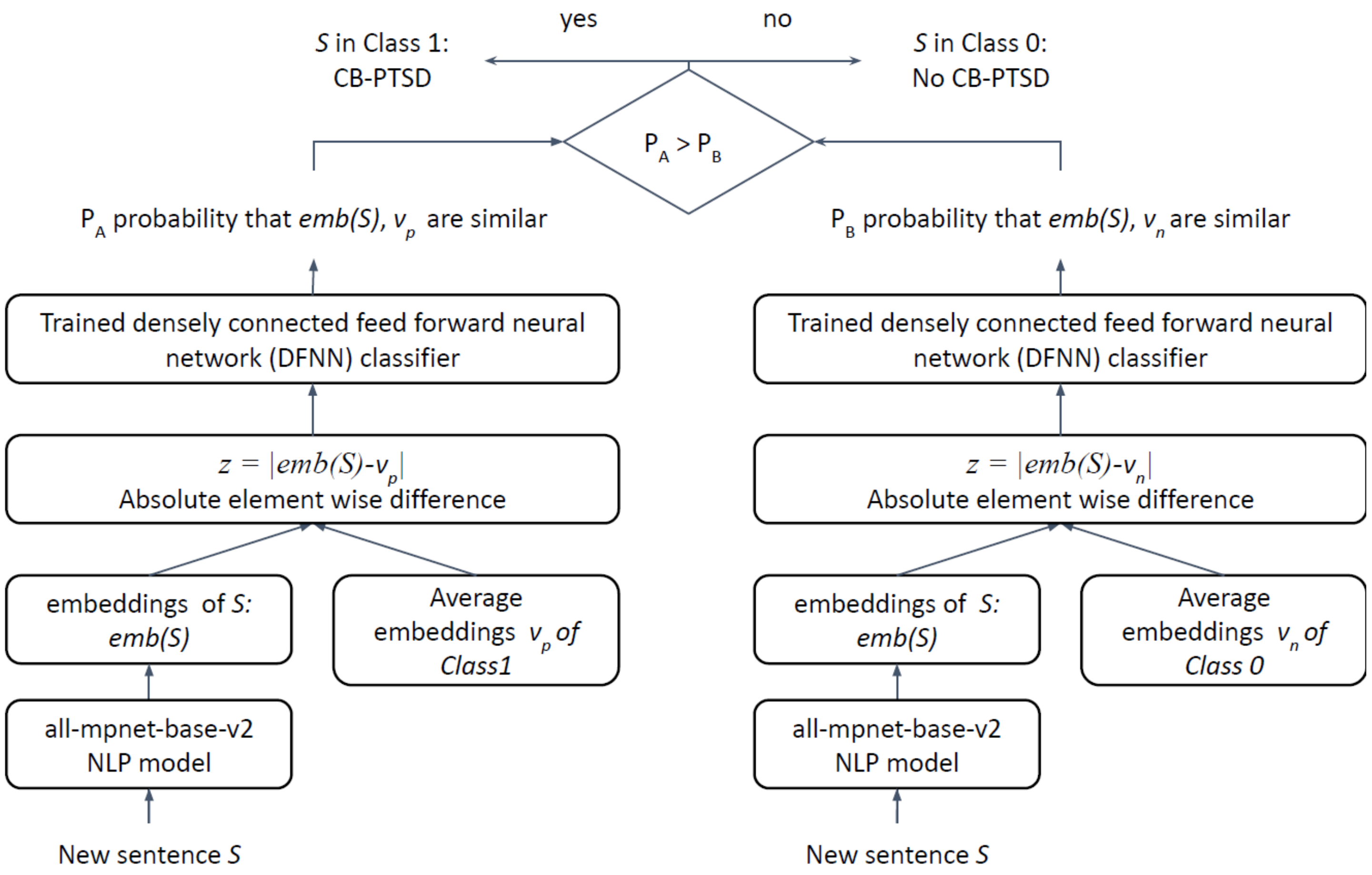


Supplemental Figure 2. Identification of childbirth-related post-traumatic stress disorder (CB-PTSD) using the developed model by classification of a new narrative sentence ($S$) that was not used for model training.

**Appendix B: Sensitivity Analysis**

To find the best parameters for the CB-PTSD classification model, the following parameter values were tested:

(1) **Model training.** We examined train/test splits of 70/30%, 75/25%, 85/15%, and 90/10%. The best results were achieved for 75/25% split.

(2) **Narrative embeddings.** To find the best narrative embeddings, we tested the following 5 pre-trained Sentence-Transformers models: `all-mpnet-base-v2`, `multi-qa-mpnet-base-dot-v1`, `all-distilroberta-v1`, `all-MiniLM-L12-v2`, and `all-MiniLM-L6-v2`. The best results were achieved for the `all-mpnet-base-v2`.

(3) **CB-PTSD classification.** We tested various PCL-5 cutoffs in the range of $[27, 33]$ and identified the cutoff of 31 as the best value for the developed model.

(4) **Neural network architectures.** We examined various neural network architectures for the DFNN model with the following parameters: number of hidden layers $[1, 3]$; hidden layer width $[12, 256]$; batch size: $[10, 256]$; optimizer: Adam; activation function: ReLU; and a Sigmoid output activation function.
